# The negative consequences of failing to communicate uncertainties during a pandemic: The case of COVID-19 vaccines

**DOI:** 10.1101/2021.02.28.21252616

**Authors:** Eleonore Batteux, Avri Bilovich, Samuel G. B. Johnson, David Tuckett

**Affiliations:** University College London; University of Warwick

**Author notes:** Corresponding author: Eleonore Batteux, Division of Psychology and Language Sciences, University College London, 1-19 Torrington Place, London, WC1E 7HB, UK.

**Keywords:** uncertainty, health communication, trust, vaccine uptake, COVID-19

## Abstract

Uncertainties pervade our health choices, particularly in the context of a novel pandemic. Despite this, rather little is known about when and how to effectively communicate these uncertainties. The focus in the medical literature so far has been on how patients respond to mentions of uncertainty relating to diagnosis or treatment, showing that these can have detrimental effects on trust and satisfaction. On the other hand, how patients are affected by these communications over time, particularly in the face of conflicting information, has received little attention. This is particularly important in the context a novel pandemic where uncertainty is rife and information changes over time. To fill this gap, we conducted an online study with UK participants on hypothetical communications relating to COVID-19 vaccines. Participants first read a vaccine announcement, which either communicated with certainty or uncertainty, and then received information which conflicted with the announcement. Those who were exposed to the certain announcement reported a greater loss of trust and vaccination intention than those who were exposed to the uncertain announcement. This shows that communicating with unwarranted certainty can backfire in the long-term, whereas communicating uncertainties can protect people from the negative impact of exposure to conflicting information.

---

No decision in healthcare comes without a degree of uncertainty. When recommending a treatment, a medical professional knows its effectiveness and possible side effects, along with their associated probabilities, what we call *risks*. But she may also be aware there is also uncertainty surrounding that probability estimate, sometimes called *ambiguity* or *radical uncertainty*. This kind of uncertainty is particularly salient in a pandemic, where we often do not know enough about the effectiveness of treatments and policies to be confident of their outcomes. In the case of COVID-19, vaccines may have been approved for use in many countries, but research is still underway to confirm their effectiveness and risks. There is even greater uncertainty relating to the impact of the vaccination programme on the pandemic more broadly. Will vaccines reduce transmission? When will restrictions be lifted? Could new variants render vaccines ineffective? These questions are full of unknown parameters.

Despite the prevalence of uncertainty, there is a lack of consensus on how best to communicate it [1]. A first step towards it has been to investigate how patients respond to communications of uncertainty, both in terms of whether they understand it and how it affects their decision-making. This work has largely painted a negative picture of uncertainty, which has led to interrogations on how best to communicate it (if at all) [2]. We take a different approach in this paper, where we investigate the negative consequences of *failing* to communicate uncertainties. Are there times where, however difficult it may be to communicate uncertainties, doing so is better than hiding them? Does failing to communicate uncertainties backfire if people find out they exist and are exposed to conflicting information? We explore these questions in the context of COVID-19 vaccines by investigating how people respond to conflicting vaccine communications.

## Communicating uncertainty in health

Before we turn to previous research on communicating uncertainty, it is worth discussing the different forms uncertainty can take. In this paper, we make a distinction between risk or probabilistic uncertainty (e.g. 20% chance of benefit from treatment) and uncertainty, or what can also be referred to as ambiguity. Uncertainty can take various forms: imprecision (e.g. 10-30% chance of benefit from treatment), conflict (e.g. experts disagreeing), lack of information (e.g. insufficient evidence) [2]. All three of these are present during a novel pandemic such as COVID-19, so we consider them together in this paper. Regarding vaccines, there is imprecision relating to their effectiveness and a lack of information on how they will affect transmission and restrictions, which may also be accompanied by conflicting opinions.

Uncertainty is communicated to varying degrees across healthcare. Physicians mention some form of uncertainty in most of their patient encounters, although this tends to be in vague terms (e.g. ‘There is a chance it will/won’t work’) [3]. They express verbal uncertainty (e.g. “I don’t know or “It’s not clear”) in most clinic visits, although more so with more educated patients [4]. In fact, physicians are less likely to communicate uncertainty if they believe patients will have negative reactions to it, which is what they tend to believe [5]. Interventions designed to communicate information to patients mention uncertainty less frequently. Although they often include quantitative risk estimates, mentioning uncertainty tends to be the exception [1,6]. When they do, this usually takes the form of verbal uncertainty (e.g. “about” or “up to”) and only a minority include numerical uncertainty (e.g. confidence intervals or ranges). This highlights the lack of consensus for how and when to communicate uncertainty in health.

We need to know how communicating uncertainty affects people’s judgments and decisions in a public health context, which can be informed by research on patients. Firstly, there are concerns it reduces understanding [7]. Patients have difficulty acknowledging there are uncertainties associated to quantitative risk estimates [8]. This could be because people generally think science can provide certainty [9] and therefore interpret expressions of uncertainty as incompetence rather than an inevitable feature of science. In addition, if a range of outcomes or probabilities are given, patients place disproportionate weight on the higher end of the range, particularly when experts disagree [9,10]. Explaining why there is uncertainty might help to mitigate misunderstandings, which has been recommended when communicating uncertainty in general [11].

Secondly, uncertainty can have negative effects on patients. Verbal expressions of uncertainty by doctors can lower patient confidence [12] and satisfaction [3,13]. Accompanying these by behaviors like positive talk, partnership building and giving information can actually increase patient satisfaction [4]. Involving patients in decisions can also mitigate any negative impact on satisfaction [3], which is positive given shifts towards shared decision-making. Behavioral expressions of uncertainty such as referring to a book or computer do not lower patient confidence [12] and can also reduce the negative impact of verbal expressions of uncertainty [13]. However, behaviors such as less fluent speech, less eye contact and fidgeting can reduce trust [14]. Finally, numerical expressions of uncertainty (e.g. ranges) can have detrimental effects. Ranges can reduce trust and credibility [7,15]. They can increase perceptions of risk and worry, although less so when communicated visually compared to textually [7,8,16]. However, it is worth noting that ranges do not necessarily have detrimental effects on trust and decision-making in other domains, where the evidence is more mixed and often context-dependent [17–19]. Taken together, findings in the medical domain show that the way in which uncertainties are communicated can greatly affect how patients respond to them.

We focus on the effects of communicating uncertainty in public health, which are similar to the effects on patients but with differences worth considering. Discussing uncertainty around numerical risk estimates may decrease perceived competence but also increase perceived honesty [9,20]. Interestingly, people report preferring to see precision in communications, but would rather uncertainties be disclosed if they exist [9]. This suggests that if people are aware that uncertainties exist they may be suspicious of communications which do not mention them. A previous study of particular interest here investigated how people respond to a government official announcing a vaccine during a hypothetical novel pandemic. Those who received uncertain communications reported lower vaccination intention due to lower perceived risk of the virus and vaccine effectiveness, accompanied by lower trust in the official [21]. This is not particularly surprising as the communications they used were verbal and highly uncertain (e.g. “we are not sure exactly how effective it will be”). This is different to the COVID-19 context where we have more precise information, despite prevailing uncertainties and changing recommendations. People may also expect uncertainties and therefore welcome their disclosure.

Parallels can be drawn between public health communication and science communication more broadly. In science communication, a lack of consensus is damaging whereas scientific uncertainty, such as ranges or a lack of evidence, is not and can even have positive effects [22]. Those who perceive science as uncertain are more favorable to uncertainty, echoing findings that if people expect uncertainty they want it communicated [9]. Interestingly, those who have high trust in science more strongly support a policy as consensus between experts increases, whereas high consensus actually lowers support in those who have low trust in science, possibly because it looks like collusion [23]. This poses a challenge to public health communication during a crisis, where addresses to the nation can be less personalized than during physician consultations for example. It is also worth noting that people use their own beliefs to interpret uncertain information, creating the risk of motivated reasoning, where people see what they want to see [22,24]. This would be problematic in a context where public opinion is divided and conspiracy theories pervasive as uncertainty can fuel those beliefs, although perhaps communicating with certainty entrenches them even further.

## What if uncertainties are not communicated?

In this study, we take a novel angle that has not been addressed in the existing literature. Are there negative consequences of *not* communicating uncertainties? When uncertainties do exist, can ignoring them backfire and eventually lead to worse outcomes? The existing literature indicates there are advantages to not communicating uncertainties, but it does not address the consequences once people are confronted with information which seems to conflict with what they were communicated. There are many instances where this applies. If a physician tells a patient their risk of developing an illness is 10% and the patient later develops it, do they lose trust in the physician? If the physician had discussed uncertainties surrounding that estimate, could that have mitigated a loss of trust? The same applies to a public health context, where a vaccine might be 90% effective against a virus but that does not mean the vaccinated are certain they will not get infected. Crucially, in contexts where evidence is lacking, new evidence can arise which invalidates previous communications. This could have detrimental effects which perhaps could be attenuated by being clear from the outset on the quality of evidence. Although disclosing uncertainties might have negative effects initially, over time it could protect against the consequences of people experiencing undesirable outcomes or conflicting information, which we know is particularly damaging in science communication [22].

This question has been explored in other contexts, where findings tend to suggest that communicating uncertainty can be beneficial in the long term. In an intelligence context, when people are told a terrorist attack occurred and shown the forecasts, they find forecasters who communicated with ranges as more credible and less worthy of blame than those who communicated point estimates [25]. In a geological context, there is no evidence of a difference between point and range forecasts in terms of perceived correctness and loss of credibility after unlikely events occur [19]. In a financial investment context, when forecasts of future returns turn out to be incorrect, forecasters who communicated with confidence and precision are perceived as less trustworthy than those who acknowledged uncertainty [26]. Interestingly, this did not lead investors to lose confidence in and pull out of their investment, showing that they blame the forecaster for incorrect forecasts but not the object of the forecast. It is worth investigating whether this applies to a medical context, i.e. whether failing to communicate uncertainties has worse consequences for confidence in the communicator than in the object of the communication (e.g. a treatment or vaccine).

## The present research

In this paper, we examine how uncertain communications affect trust and vaccination intention over time. Specifically, we test whether communicating uncertainty about COVID-19 vaccines limits any loss of trust and vaccination intention after people receive conflicting information about their effectiveness. We focus on COVID-19 for two reasons. The first being that we urgently need to understand how to effectively communicate about COVID-19 vaccines to maximize uptake and ensure the successful rollout of the vaccination programme. Vaccine hesitancy is a particular concern, linked to a lack of trust [27]. Secondly, although we use hypothetical communications in our study, COVID-19 provides a real pandemic context that participants can relate to and have knowledge of. This differs from a previous study on communicating vaccine uncertainty, which referred to a hypothetical virus participants had very little knowledge of [21]. Our hypotheses were preregistered on the Open Science Framework (OSF; https://osf.io/c73px/) and are as follows.

### Hypothesis 1

We expect people are less favorable to getting vaccinated after receiving uncertain compared to certain vaccine communications. This is in keeping with the literature on how people respond to uncertain communications in health and public health. Our primary outcome variable is vaccination intention, which we expect to be lower following uncertain communications as found in a previous study [21]. We investigate whether this is accompanied by lower perceptions of vaccine effectiveness, as has been found previously [21], stronger avoidance emotions (e.g. worry) and weaker approach emotions (e.g. excitement). Indeed, we expect emotions to be crucial to people’s decision-making in contexts of uncertainty [28]. Our second key interest is how uncertainty affects trust in communicators, which is crucial to both vaccine uptake and compliance to guidelines during a pandemic [27,29]. Previous studies suggest trust should be lower [15,21].

### Hypothesis 2

Once people receive information which conflicts with earlier communications, we expect those who initially received certain communications experience more negative effects compared to those who received uncertain communications. We posit that communicating uncertainty makes people more likely to expect information to change over time and therefore less surprised and disappointed when confronted to new and conflicting information, as has been found in the financial domain [26]. On the other hand, communicating with unwarranted certainty may be perceived as intentionally misleading. This would not be surprising in the context of COVID-19 in the UK where government overpromising has eroded trust [30]. We expect to see greater reductions in vaccine intention in those receiving certain communications, accompanied by greater reductions in trust, perceived vaccine effectiveness and approach emotions and a greater increase in avoidance emotions.

To test these hypotheses, we conducted a study in November 2020, before any COVID-19 vaccine announcement and effectiveness rates were widely communicated. We presented participants from the general UK population with a hypothetical vaccine announcement containing information about the vaccine’s effectiveness. Some received information which conveyed certainty about the rate of effectiveness whereas others received information which conveyed uncertainty. Participants were then told that they find some new research on the vaccine’s effectiveness, which is significantly lower than communicated in the announcement for both the certain and uncertain announcement. We compare participants’ vaccination intention, trust, perceived vaccine effectiveness and affective reactions after receiving the announcement to after receiving conflicting information.

## Method

### Design

Communication certainty (1-certain, 2-uncertain) was manipulated between-subjects. Participants were randomly allocated to the certain or uncertain communication.

### Participants

328 participants residing in the UK were recruited using Prolific, an online participant recruitment platform (https://www.prolific.co/). A sample of 328 was required to find a small effect (*d*=0.20) for Hypotheses 2a-e with a mixed model ANOVA with high power (>.95) and alpha level (<.05). This sample size also allows enough power to test Hypothesis 1 in accordance with existing findings. Participants were compensated for their time at a rate of £7.50 per hour. They were asked demographic questions (age, gender, level of education) and questions about COVID-19. Firstly, how much trust they currently have in the government’s handling of the COVID-19 crisis on a 5-point scale (1-not at all, 5-a great deal). Secondly, how reliable, precise and consistent they perceive the science relating to COVID-19 on a 7-point scale (1-reliable/precise/consistent, 7-unreliable/imprecise/inconsistent). These were added to provide an overall score on their perception of the certainty of COVID-19 science. Finally, participants completed the Vaccination Attitudes Examination scale which provides an overall score of favorability to vaccination [31] on a 5-point scale (1-strongly disagree, 5-strongly agree). Participant characteristics can be found in Table 1.

**Table 1:**
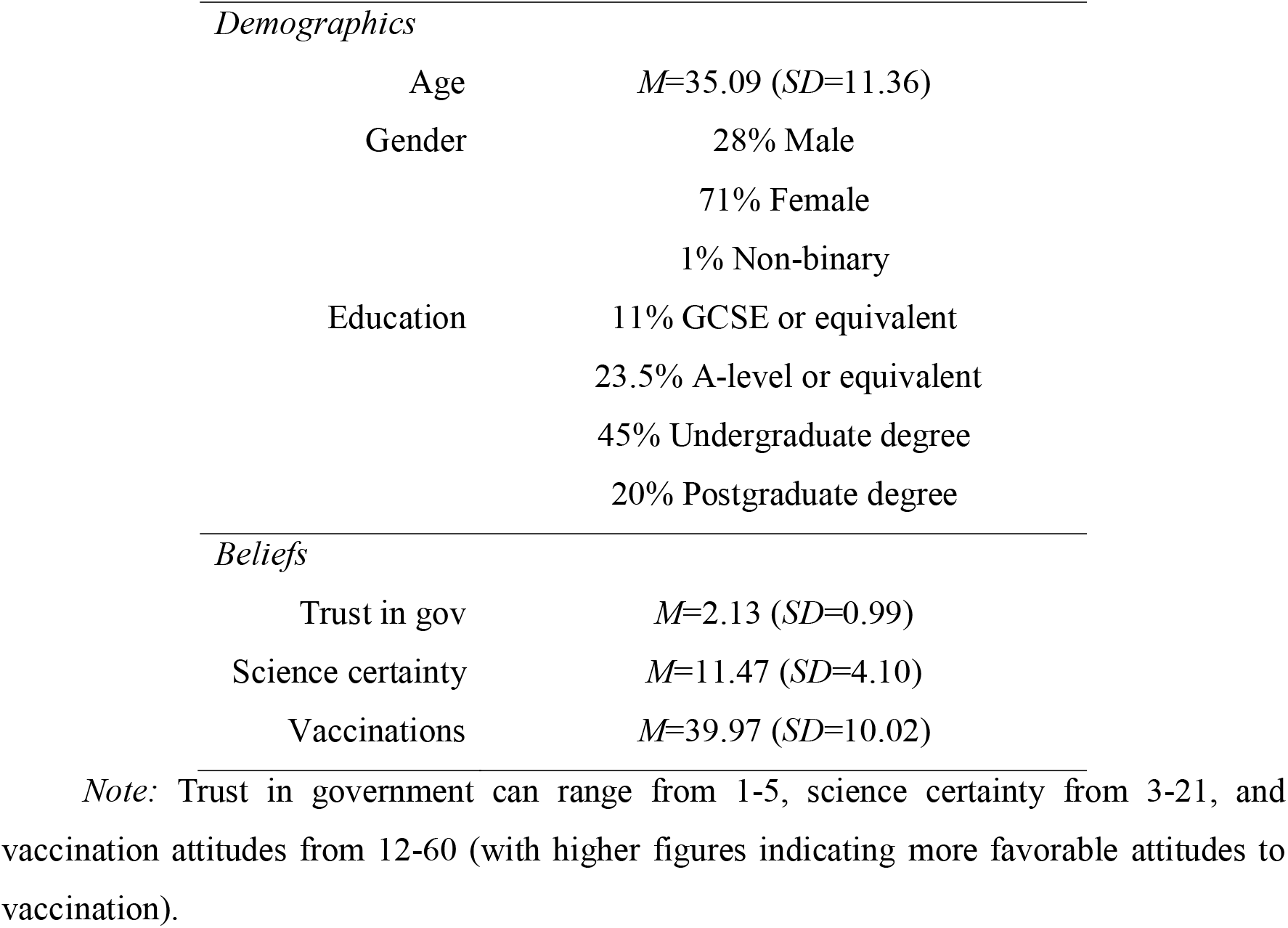
Participant characteristics

|  |  |
| --- | --- |
| Age | $M=35.09$ ($SD=11.36$) |
| Gender | 28% Male<br>71% Female<br>1% Non-binary |
| Education | 11% GCSE or equivalent<br>23.5% A-level or equivalent<br>45% Undergraduate degree<br>20% Postgraduate degree |
| *Beliefs* |  |
| Trust in gov | $M=2.13$ ($SD=0.99$) |
| Science certainty | $M=11.47$ ($SD=4.10$) |
| Vaccinations | $M=39.97$ ($SD=10.02$) |

*Note:* Trust in government can range from 1-5, science certainty from 3-21, and vaccination attitudes from 12-60 (with higher figures indicating more favorable attitudes to vaccination).

### Scenario

Participants were reminded they are in the middle of the COVID-19 pandemic and told to imagine they hear a public health government representative make a vaccine announcement on the news. This announcement states that a vaccine has passed the necessary checks and will soon be available. For those in the certain condition the representative says: “I can confirm that the vaccine is 60% effective. This means that, although the vaccine might not work for everyone, there is a very good chance that it will work for you. This vaccine will significantly drive down the infection rate and we will be able to remove the restrictive measures we put in place to combat the virus.” In the uncertain condition the representative says: “The vaccine is between 50 and 70% effective. The reason I can’t give a more precise estimate is because the data we have doesn’t allow that. There might be some things we don’t know yet about the vaccine, but this is the best available option. Although it might not work for everyone, there is a chance it will work for you. This vaccine will hopefully drive down the infection rate and we may be able to remove the restrictive measures we put in place to combat the virus.” Then, all participants are told: “a week later, the vaccine is available and you can book an appointment with your local GP practice. Before deciding whether to get it, you want to read the research on the vaccine’s effectiveness. You find the latest international piece of research which is deemed to have the most reliable data. This tells you that the vaccine is actually nearer to 40% effective.”

### Measures

Measures were taken after participants received the initial announcement and after they read the additional research about the vaccine’s effectiveness. Participants were asked how much confidence and trust they have in the government representative, how effective they think the vaccine is, how they feel about getting the vaccine (excited, confident, worried, uncertain) on 5-point scales (1-not at all, 5-a great deal) and how likely they are to get the vaccine on a 5-point scale (1-definitely not, 5-definitely yes).

## Results

Our findings are broadly consistent across measures of vaccination intention, vaccine effectiveness, trust and confidence in government and emotion. They support the hypothesis that conflicting information leads to more negative effects among those who were exposed to certain compared to uncertain communications (Hypothesis 2). However, they do not support the hypothesis that people are initially more favorable to certain compared to uncertain communications (Hypothesis 1). The data can be found on OSF (https://osf.io/c73px/).

### Vaccination

As expected, the certain announcement led to a greater decline in vaccination intention following exposure to conflicting information (see Figure 1). Indeed, there was no difference in vaccination intention between people who received the certain and uncertain announcement after the announcement (*t*_326_=-0.12, *p*=.903, *d=*0.01), but there was a marginal difference after reading the conflicting information (*t*_326_=-1.804, *p*=.072, *d=*0.20) (*F*_1,326_=9.50, *p*=.002, *η*_*p*_^*2*^=0.03). In other words, those who received the certain announcement experienced a greater reduction in vaccination intention than those who received the uncertain announcement (*t*_326_=3.08, *p*=.002, *d=*0.34). Participants had stronger vaccination intentions after the announcement than after reading conflicting information (*F*_1,326_=134.47, *p*<.001, *η*_*p*_^*2*^=0.29) and there was no overall difference between those receiving the certain and uncertain announcement (*F*_1,326_=1.02, *p*=.314, *η*_*p*_^*2*^<0.01).

**Figure 1:**
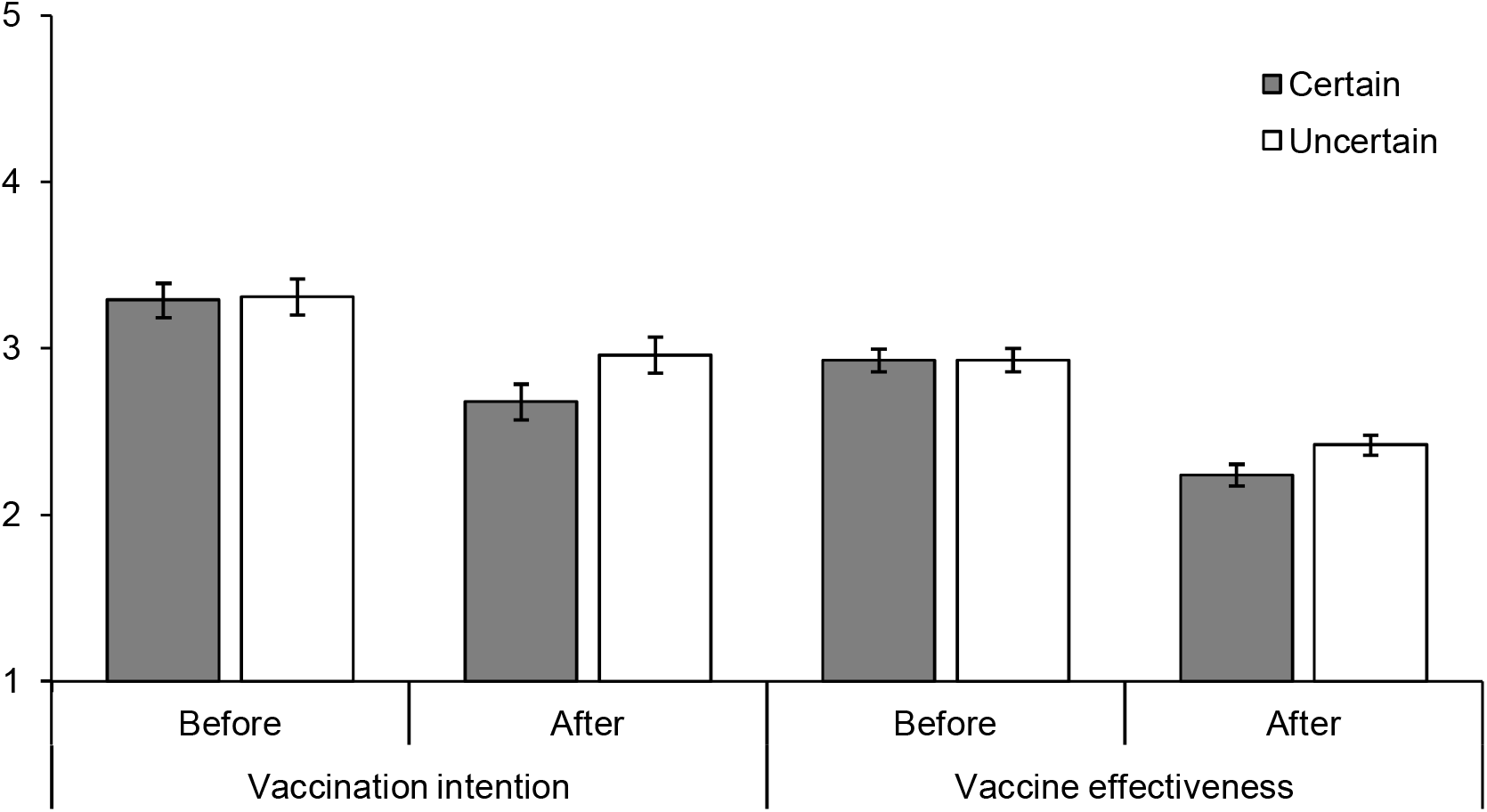
Vaccination intention and perceived vaccine effectiveness before receiving conflicting information (i.e. after the vaccine announcement) and after receiving conflicting information by announcement certainty.

The pattern of findings was the same for effectiveness, where the certain announcement led to a greater decline in perceived effectiveness (see Figure 1). Participants receiving the certain and uncertain announcement perceived the effectiveness equally after the announcement (*t*_326_=0.06, *p*=.951, *d=*0.01), whereas those who received the certain announcement perceived it as less effective after reading conflicting information (*t*_326_=-1.99, *p*=.048, *d=*0.22) (*F*_1,326_=5.45, *p*=.020, *η*_*p*_^*2*^=0.02). Participants thought the vaccine was more effective after the announcement than after reading conflicting information (*F*_1,326_=232.63, *p*<.001, *η*_*p*_^*2*^=0.42) and there was no overall significant difference between those receiving the certain and uncertain announcement (*F*_1,326_=1, *p*=.318, *η*_*p*_^*2*^<0.01).

### Government

The certain announcement led to a greater decline in trust and confidence in the government official after exposure to conflicting information (see Figure 2). Both groups were equally trusting of the government official after the announcement (*t*_326_=-0.54, *p*=.957, *d=*0.01), whereas those who received the certain announcement were less trusting after reading conflicting information (*t*_326_=-3.04, *p*=.003, *d=*0.34) (*F*_1,326_=9.54, *p*=.002, *η*_*p*_^*2*^=0.03). Participants had more trust in the government official after their announcement than after reading conflicting information (*F*_1,326_=187.12, *p*<.001, *η*_*p*_^*2*^=0.37) and there was no overall significant difference between those receiving the certain and uncertain announcement (*F*_1,326_=2.70, *p*=.101, *η*_*p*_^*2*^=0.01).

**Figure 2:**
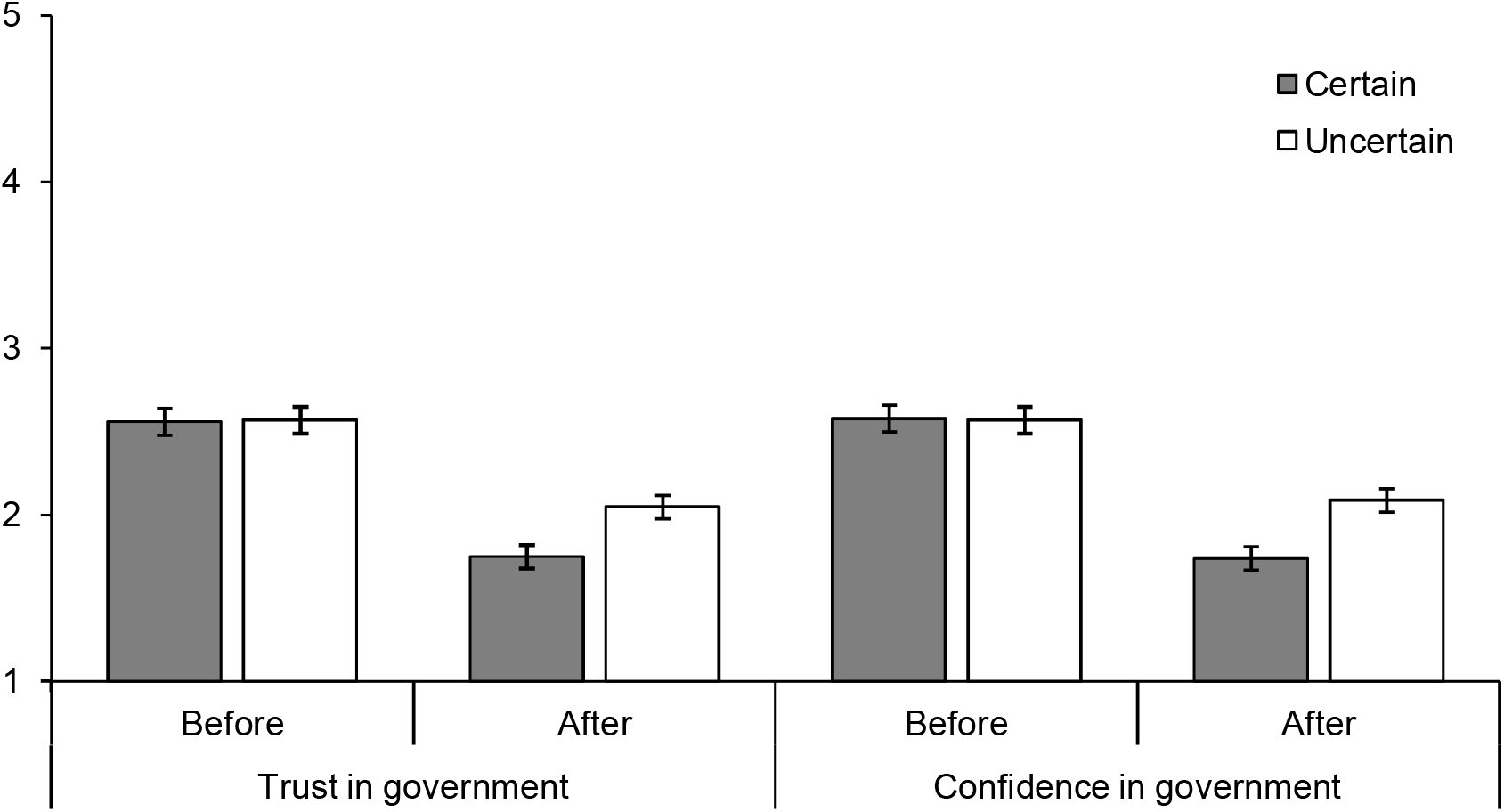
Trust and confidence in the government official who made the vaccine announcement before receiving conflicting information (i.e. after the vaccine announcement) and after receiving conflicting information by announcement certainty.

This was also the case for confidence (see Figure 2). Both groups were equally confident in the government official after the announcement (*t*_326_=0.79, *p*=.914, *d=*0.01), whereas those who received the certain announcement were less confident after reading conflicting information (*t*_326_=-3.45, *p*=.001, *d=*0.38) (*F*_1,326_=12.08, *p*=.001, *η*_*p*_^*2*^=0.04). Participants were more confident in the government official after their announcement than after reading conflicting information (*F*_1,326_=170.61, *p*<.001, *η*_*p*_^*2*^=0.34) and there was no overall significant difference between those receiving the certain and uncertain announcement (*F*_1,326_=3.13, *p*=.078, *η*_*p*_^*2*^=0.01).

### Predictors of vaccination intention

In a previous study on communicating uncertainty about vaccines during a pandemic, perceived vaccine effectiveness mediated the relationship between communicated uncertainty and vaccination intention but trust in the government official did not [21]. We explored whether this was also the case here using the PROCESS macro for SPSS [32] (see Figure 3). We find that both trust in the government official (*b*=0.09, 95% CI[0.02,0.18]) and perceived effectiveness (*b*=0.14, 95% CI[0.003,0.29]) mediate the relationship between announcement certainty and vaccination intention. This means that participants who received the uncertain announcement were more likely to want to get vaccinated, both because they had higher trust in the government official and because they perceived the vaccine as more effective after receiving conflicting information. Both of these mechanisms contribute to the effect of uncertainty communication on vaccination intention. Trust may not explain the effect of uncertainty communication on vaccination intention when the announcement is made [21], but it does here after participants are exposed to conflicting information.

**Figure 3:**
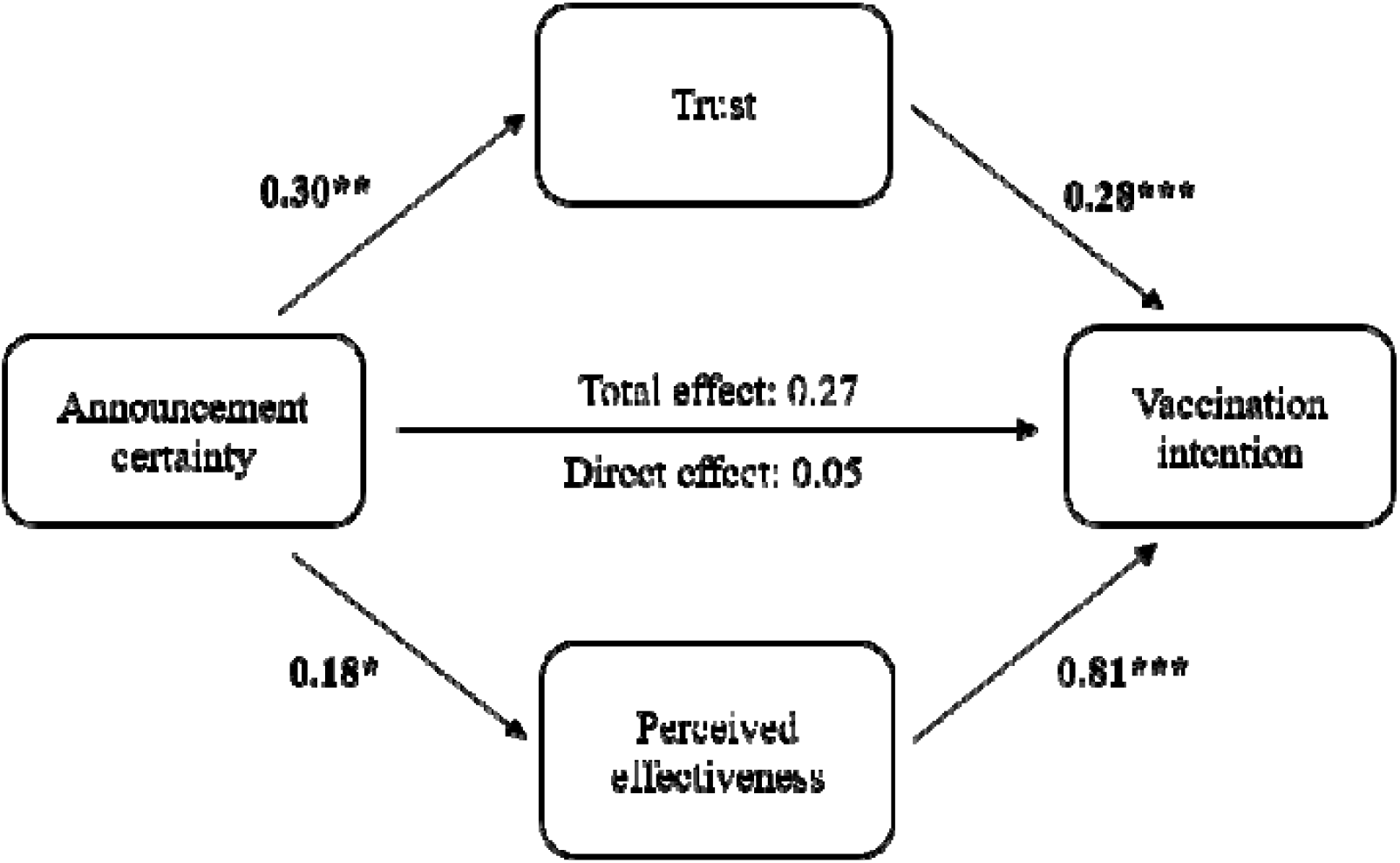
Relationship between announcement certainty and vaccination intention after receiving conflicting information mediated by trust in government official and perceived vaccine effectiveness. * refers to *p*<.05, ** refers to *p*<.01, *** refers to *p*<.001.

### Emotions

Although the pattern of findings on emotions is similar, the differences between those receiving the certain and uncertain announcement were less clear, perhaps due to the hypothetical nature of the study. The certain announcement led to a greater increase in avoidance emotions after exposure to conflicting information (see Figure 4). Participants were less worried after the announcement than after reading conflicting information (*F*_1,326_=60.50, *p*<.001, *η*_*p*_^*2*^=0.16), which was qualified by an interaction with the certainty of the announcement (*F*_1,326_=4.86, *p*=.028, *η*_*p*_^*2*^=0.02). Those who received the certain announcement experienced a greater increase in worry than those who received the uncertain announcement (*t*_326_=-2.20, *p*=.028, *d=*0.24), although there was no statistical difference between each group after receiving the announcement (*t*_326_=-0.97, *p*=.332, *d=*0.11) or reading the conflicting information (*t*_326_=0.51, *p*=.614, *d=*0.06). There was no overall significant difference between those receiving the certain and uncertain announcement (*F*_1,326_=0.05, *p*=.819, *η*_*p*_^*2*^<0.01). Participants were less uncertain after the announcement than after reading conflicting information (*F*_1,326_=19.35, *p*<.001, *η*_*p*_^*2*^=0.06), which was qualified by an interaction with the certainty of the announcement (*F*_1,326_=9.27, *p*=.003, *η*_*p*_^*2*^=0.03). Those who received the certain announcement experienced a greater increase in uncertainty than those who received the uncertain announcement (*t*_326_=-3.05, *p*=.003, *d=*0.34), although there was no statistical difference between each group after receiving the announcement (*t*_326_=-1.70, *p*=.091, *d=*0.19) or reading the conflicting information (*t*_326_=0.74, *p*=.462, *d=*0.08). There was no overall significant difference between those receiving the certain and uncertain announcement (*F*_1,326_=0.24, *p*=.628, *η*_*p*_^*2*^<0.01).

**Figure 4:**
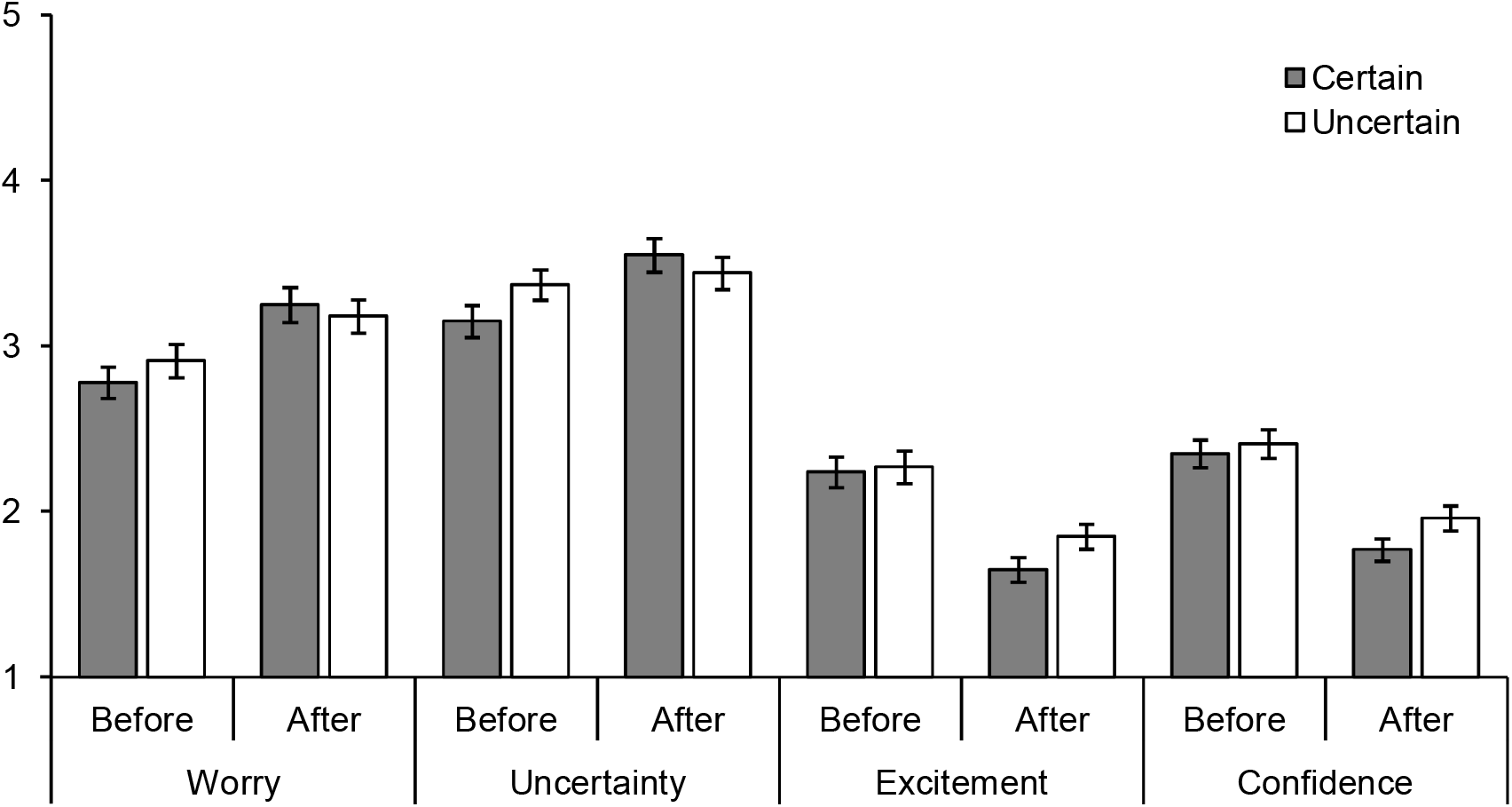
Emotions before receiving conflicting information (i.e. after the vaccine announcement) and after receiving conflicting information by announcement certainty.

We do not find that the certain announcement leads to a greater decrease in approach emotions after conflicting information (see Figure 4). Participants were more excited about the vaccine after the announcement than after reading conflicting information (*F*_1,326_=127.76, *p*<.001, *η*_*p*_^*2*^=0.28) but the interaction with the certainty of the announcement was marginally significant (*F*_1,326_=1.20, *p*=.060, *η*_*p*_^*2*^=0.01). There was no overall significant difference between those receiving the certain and uncertain announcement (*F*_1,326_=1.05, *p*=.306,*pη* ;_*p*_ ^*2*^<0.01). Participants were more confident about the vaccine after the announcement than after reading conflicting information (*F*_1,326_=126.09, *p*<.001, *η*_*p*_^*2*^=0.28) but the interaction with the certainty of the announcement was not significant (*F*_1,326_=2.16, *p*=.142, *η*_*p*_^*2*^=0.01). There was no overall difference between those receiving the certain and uncertain announcement (*F*_1,326_=1.41, *p*=.235, *η*_*p*_^*2*^<0.01).

## Discussion

Communicating uncertainties had protective effects against new conflicting information. Participants showed a greater reduction in vaccination intention after receiving information which conflicted with communications delivered with certainty, as opposed to communications which acknowledged uncertainties. This was accompanied by a greater reduction in trust in the communicator and perceived vaccine effectiveness, which both affected vaccination intention. Participants also experienced a greater increase in avoidance emotions (worry and uncertainty) following information which conflicted with certain as opposed to uncertain communications. We did not find a decline in approach emotions, although they were quite low to begin with.

The picture is more complicated when it comes to differences between certain and uncertain communications at each time point. At the time of the vaccine announcement, we do not find clear evidence that those who received uncertain communications are less likely to get vaccinated. This stands in contrast to previous findings, although communications in those studies expressed greater uncertainty than in ours [21]. Although most of the previous literature indicates that communicating uncertainty has damaging effects [2], our findings are an example of the kinds of contexts in which those effects might be weaker, i.e. a context where uncertainty is particularly salient. Although patients might not expect scientific uncertainty generally [9], people have been exposed to it during COVID-19 and may therefore expect it and want it to be communicated [22].

Once people receive information which conflicts with the vaccine announcement, we start to see differences between those exposed to the certain and uncertain announcement. The crucial difference is that the government official who delivered the announcement appears more trustworthy to those who were exposed to uncertainty. Communicating with unwarranted certainty damages trust, which echoes the finding that the UK government’s overpromising during the COVID-19 pandemic eroded trust [30]. Although those who received the certain announcement now perceive the vaccine as less effective, the difference with vaccination intention is less clear. Having said that, those who experience a strong decline in trust and perceived vaccine effectiveness following the certain announcement also experience a strong decline in vaccination intention, making it weaker compared to those who received the uncertain announcement. Although communicating with certainty about vaccines is more damaging for trust in communicators than for vaccination intention, as findings in the financial domain suggested [26], the effects on vaccination intention remain a problem.

### Limitations

Our findings highlight the benefits of communicating uncertainties in health, but they are only a starting point. Given their limitations, we recommend caution in interpreting and implementing these findings. We largely focused on uncertainties relating to vaccine effectiveness. Figures of vaccine effectiveness are often communicated with precision (e.g. 70% effectiveness) even though they come with confidence intervals, which we communicated in this study. Beyond this, there are many uncertainties relating to vaccines during a novel pandemic worth exploring. Risks of side effects, including unforeseen risks which are not detectable in trials over short time periods, are particularly important to the public when making vaccination decisions [33]. Many are motivated to get vaccinated to reduce the spread of the virus and lift restrictions in place to control it, although whether the vaccination programme can do so is not necessarily known until well underway [34]. It is unclear which of these uncertainties will have a stronger impact on vaccination intention, although we expect exposure to conflicting information relating to all of these to have similar effects to those we report here. We also do not know what the combined effects are of all these uncertainties and multiple exposures to conflicting information. We only exposed participants to one instance of conflicting information, whereas there are likely to be many more throughout a pandemic. Vaccination intention and particularly trust are likely to evolve over time and may be more impacted by repeated exposures.

Given the hypothetical nature of our study, caution is warranting when applying findings. Although we used hypothetical communications, we focused on a real pandemic situation where participants had prior knowledge and experiences relating to COVID-19. Participants are likely to have been more engaged and invested in the study than in completely hypothetical studies. However, we used a hypothetical delay between the vaccine announcement and receiving conflicting information. This makes generalization to real instances more difficult, given that time delays increase the likelihood that people forget the information they receive and therefore do not interpret new information as conflicting with it. Having said that, government communications and new information are likely to be highly mediatized and conflicts made salient during a crisis, as was the case during COVID-19.

Finally, it would be valuable to know how well these findings generalize beyond a pandemic context in the UK. Indeed, it is worth investigating whether our findings generalize to other situations, such as physician-patient interactions where communicating uncertainty seems initially problematic but may have long-term benefits that have not been uncovered yet. In addition, generalizing beyond the UK context would be valuable to inform global communication practices. Given that trust in government is important for vaccine uptake beyond the UK [27], we expect findings would be similar in other countries.

### Implications for communicating uncertainty in health

Our findings support uncertainties being communicated in healthcare by highlighting the negative consequences of failing to do so. Even though communicating with certainty can initially have benefits, if that certainty is not warranted this can have damaging consequences in the long run. When considering whether to disclose uncertainties, communicators should consider the quality of the evidence and whether people are likely to be exposed to diverging opinions and conflicting information. Anticipating this by discussing uncertainties could avoid negative consequences further down the line. In contexts of great uncertainty, people may not actually be averse to uncertainties being communicated, unlike what previous studies in more certain contexts suggest [2]. More work is needed to establish whether people respond differently to uncertain communications depending on the level of contextual uncertainty.

Another key question is how to communicate uncertainties. Previous studies have suggested some formats are more effective, such as visual depictions of uncertainty causing less worry than verbal ones [8]. We used several ways of communicating uncertainty here, which at present we cannot tease apart. We manipulated the uncertainty of vaccine effectiveness, which was a point estimate in the certain announcement and a range in the uncertain announcement. Ranges may communicate uncertainty but they also increase worry and reduce understanding [7], suggesting that they alone are not sufficient. We accompanied the range by an explanation for the uncertainty, which could have perhaps enabled people to understand the uncertainty better. We also included verbal descriptions of uncertainty regarding the broader risks and benefits of the vaccine which may have increased people’s perception of uncertainty, perhaps making them respond less negatively to conflicting information later on. These various ways of communicating uncertainty might have contributed differently to our findings. Future research should seek to isolate their effects to better understand their relative effectiveness.

Who is best placed to communicate these uncertainties? Our study does not address this question, although we provide the following reflections which could inform future research. People might have different expectations of government compared to medical practitioners. In fact, people have particularly low levels of trust in politicians [35] and perhaps expect to be misled by government. It is conceivable that the effects we find on trust are due to participants perceiving the government official as intentionally trying to mislead them into getting vaccinated, which might not have been the case if the information came from a medical practitioner. On the other hand, people might have higher expectations of medical practitioners. They might expect certainty in their communications, thereby reacting negatively to expressions of uncertainty (although may react even more negatively if uncertainty that was not initially communicated is later revealed). Uncertainty could perhaps be interpreted as incompetence from medical practitioners but honesty from politicians, who have had a tendency to overpromise COVID-19 [30]. There may be instances where governments are better placed to communicate uncertainty, particularly during a national crisis, which further research should clarify.

### Conclusion

In a novel pandemic context, where evidence is lacking and evolves over time, people are often faced with changing and conflicting information. Under these circumstances, we show that communicating uncertainties attenuates the negative consequences of being faced with conflicting information, the most damaging form of uncertainty in science communication [22]. Although it might come with challenges, communicating uncertainty in healthcare can be beneficial for maintaining trust and patient commitment over time. It takes more account of the potential for health care communications to develop active expertise in its recipients, thereby developing shared and resilient understanding [36,37]. Our findings support calls for greater transparency and acknowledgements of uncertainty in communications relating to COVID-19 [38,39]. They highlight the advantages of communicating uncertainty, which we hope will further motivate research on doing so effectively.

## Data Availability

The data can be found on Open Science Framework.

https://osf.io/c73px/

## Acknowledgment

This research has been supported by the Think Forward Initiative (a partnership between ING Bank, Deloitte, Dell Technologies, Amazon Web Services, IBM, and the Center for Economic Policy Research – CEPR). The views and opinions expressed in this paper are solely those of the authors and do not necessarily reflect the official policy or position of the Think Forward Initiative or any of its partners.

